# COVID-related Excess Missed HIV Diagnoses in the United States in 2021: Follow-up to 2020

**DOI:** 10.1101/2023.09.28.23296285

**Authors:** Alex Viguerie, Ruiguang Song, Anna Satcher Johnson, Cynthia M. Lyles, Angela Hernandez, Paul G. Farnham

## Abstract

**Objective:** COVID-19 and related disruptions led to a significant drop in HIV diagnoses in the US in 2020. Recent analyses found 18% fewer diagnoses than expected among persons with HIV (PWH) acquiring infection in 2019 or earlier, suggesting that the drop in diagnoses cannot be attributed solely to decreased transmission. This analysis evaluates the progress made towards closing the 2020 diagnosis deficit in 2021.

**Methods:** We apply modified versions of previously developed methods analyzing 2021 diagnosis data from the National HIV Surveillance System to determine whether the 2021 diagnosis levels of PWH infected pre-2020 are above or below the projected pre-COVID trends. We apply these analyses on stratifications based on assigned sex at birth, transmission group, geographic region, and race/ethnicity.

**Results:** In 2021, HIV diagnoses returned to pre-COVID levels among all PWH acquiring infection 2011-19. Among Hispanic/Latino PWH and males, diagnoses returned to pre-COVID levels. White PWH, men who have sex with men, and PWH living in the south and northeast showed higher-than-expected levels of diagnosis in 2021. For the remaining populations, there were fewer HIV diagnoses in 2021 than expected.

**Conclusions:** While overall diagnoses returned to pre-COVID levels, the large diagnosis gap observed in 2020 remained unclosed at the end of 2021. Lower than expected diagnosis levels among certain populations indicates that COVID-19 related disruptions to HIV diagnosis trends were present in 2021. Although some groups showed higher-than-projected levels of diagnoses, such increases were smaller than the corresponding 2020 decreases. Expanded testing programs designed to close these gaps are essential.

## Background

The COVID-19 pandemic resulted in a 17% drop in HIV diagnoses in the United States in 2020, as testing and other services were severely disrupted [1, 2]. There has been uncertainty, however, in the interpretation of these data-in particular, what portion of the drop is attributable to reduced testing services versus a drop in transmission levels. A recent analysis attempted to clarify this issue by considering diagnoses only among persons with HIV (PWH) acquiring HIV in 2019 or earlier [3]. Year of infection was estimated by applying the CD4-depletion model to CD4 measurements taken at diagnosis. Such measurements form the basis for modern incidence estimation methods [4].

The analysis in [3] found that diagnoses decreased by approximately 17-18% among PWH who acquired infection in 2019 or earlier, and hence this drop cannot be explained by changes in incidence levels. The drop appeared to be uniform across estimated infection years, with diagnoses being missed among older and more recent infections at similar rates.

Females (at birth), persons who inject drugs (PWID), Hispanic/Latino PWH, and heterosexual persons missed diagnoses at higher rates as compared to other subpopulations. In total, among PWH acquiring infection in 2010-2019, an estimated 3100-3300 fewer infections than expected were diagnosed in 2020, compared to expected diagnosis levels given pre-COVID trends.

In this brief report, we perform a follow-up to the analysis in [3] for the year 2021 to assess whether the changes observed in HIV diagnosis patterns in 2020 persisted in 2021 and if the diagnosis deficit that was observed in 2020 has remained. Using modified versions of the methods developed for the 2020 analysis, we project the expected number of diagnoses in 2021, for each infection year, among PWH who acquired infection from 2011-2019. As in the 2020 analysis, we then compare these diagnosis counts with observed diagnoses levels. Analyses are performed for both the entire United States population, as well as stratifications by assigned sex at birth, transmission group, race/ethnicity, and region.

## Methods

### Diagnosis Projection

We apply modified versions of the incidence-based and diagnosis delay-based methods first introduced in [3] to project diagnoses in 2021 among PWH acquiring infection in the years 2011-2019. These methods project the expected number of diagnoses in year *y* among infections acquired in year *x* as:

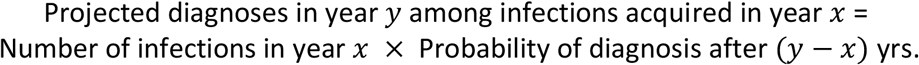

The methods differ in exactly how the number of infections in year *x* and the probability of diagnosis after (*y* − *x*) years are estimated. The mathematical details regarding differences in derivation can be found in [3] (Appendix A).

We modified the methods from [3], as the window of projection is no longer 1 year (as in the 2020 analysis), by extending it to two years. Accordingly, a validation analysis (not reported for purposes of brevity) was performed against 2019 diagnoses using data from 2011-2017 to confirm consistency with surveillance data.

Diagnoses data from the National HIV Surveillance System data for 2021 were used to perform these analyses. We note these data differ slightly from those used in [3], due to routine annual updating of NHSS data, and are consistent with those reported in [5]. Accordingly, the numbers reported here for 2020 may differ slightly from those presented in [3], which were based on the data reported in [2]. However, the overall findings and conclusions of [3] have not changed..

## Results

### Missed and Recovered Diagnoses in 2021

15,609 HIV diagnoses were observed in 2021 among PWH acquiring infection in the years 2011-19, compared with an expected total of 15,587 diagnoses (Figure 1, left), or approximately 20 more diagnoses than expected. As infections among this population were acquired prior to the onset of the COVID-19 pandemic, they are unaffected by any confounding in HIV diagnoses caused by COVID-19 related changes in incidence. These findings therefore suggest that HIV diagnoses in the United States resumed pre-COVID trends in 2021, at least among infections prior to 2020. We note further that the different methods were in good agreement with both each other and surveillance data, with the discrepancy in overall diagnoses around 1%, and the year-by-year trend discrepancy under 4%, as can be observed in Fig 1 (right).

**Fig 1:**
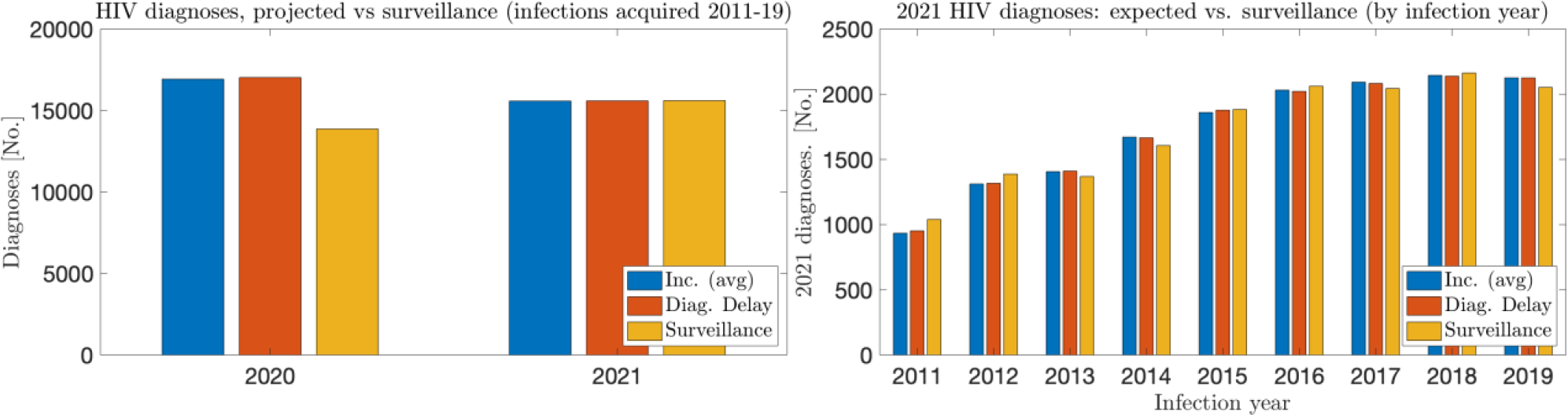
Left: Total HIV diagnoses, observed vs expected, in 2020 and 2021; Right: 2021 HIV diagnoses, observed vs expected, by infection-year.

In 2020, over the infection years 2011-19, 13,868 HIV diagnoses were observed compared to 16,978 expected diagnoses, a deficit of approximately 3,100 (Table 1, Figure 1). While diagnoses in this group returned levels consistent with pre-COVID trends in 2021, a significant cumulative deficit of ∼3090 diagnoses remains (Table 1).

**Table 1:** Total missed diagnoses (projected diagnoses minus observed diagnoses) in 2020 and 2021, and deficit at the end of 2021, by subpopulation acquiring infection 2011-2019. Numbers reported are averages of 2 projection methods. Note a negative number means more diagnoses than expected. ^1^Men who have sex with men (MSM) ^2^Heterosexual persons (HET) ^3^Persons who inject drugs (PWID) ^4^Men who have sex with men and inject drugs (MSM/PWID)

| Group | Diagnoses, 2020 |  |  |  | Diagnoses, 2021 |  |  |  | Diagnosis deficit (2020 – 21) |
| --- | --- | --- | --- | --- | --- | --- | --- | --- | --- |
|  | Expected | Observed | Missed | %missed | Expected | Observed | Missed | %missed | Total |
| Total | 16,978 | 13,868 | ~3100 | 18.3% | 15,587 | 15,609 | ~-20 | -0.2% | ~3090 |
| Black/<br>African<br>American | 7,250 | 6,055 | ~1200 | 16.5% | 6,643 | 6,472 | ~170 | 2.6% | ~1370 |
| Hispanic/<br>Latino | 4,874 | 3,187 | ~1100 | 21.7% | 4,592 | 4,567 | ~30 | 0.5% | ~1130 |
| White | 3,831 | 3,230 | ~600 | 15.7% | 3,437 | 3,552 | ~-120 | -3.4% | ~480 |
| Male (at<br>birth) | 14,164 | 11,764 | ~2400 | 16.9% | 13,069 | 12,991 | ~80 | 0.6% | ~2480 |
| Female (at<br>birth) | 2,822 | 2,170 | ~650 | 23.1% | 2,535 | 2,439 | ~100 | 3.8% | ~750 |
| Northeast | 2,388 | 1,921 | ~500 | 19.6% | 2,175 | 2,197 | ~-20 | -1.0% | ~480 |
| Midwest | 2,240 | 1,884 | ~350 | 15.9% | 2,089 | 1,997 | ~100 | 4.4% | ~450 |
| West | 3,555 | 2,925 | ~650 | 17.7% | 3,275 | 3,055 | ~220 | 6.7% | ~870 |
| South | 8,804 | 7,203 | ~1600 | 18.1% | 8,075 | 8,179 | ~-100 | -1.3% | ~1500 |
| MSM<sup>1</sup> | 11,852 | 9,944 | ~1900 | 16.1% | 10,942 | 10,991 | ~-50 | -0.4% | ~1850 |
| HET<sup>2</sup> | 3,182 | 2,391 | ~800 | 24.9% | 2,892 | 2,792 | ~100 | 3.5% | ~900 |
| PWID<sup>3</sup> | 1,303 | 1,054 | ~250 | 19.1% | 1,194 | 1,153 | ~40 | 3.4% | ~290 |
| MSM/<br>PWID<sup>4</sup> | 655 | 459 | ~200 | 31.3% | 594 | 522 | ~70 | 12.1% | ~270 |

Diagnosis levels did not recover uniformly across groups. In particular, White PWH, PWH in the South and Northeast, and men who have sex with men (MSM) showed signs of higher-than-expected diagnosis levels in 2021, indicating some progress towards closing the 2020 diagnosis deficit among those subpopulations (Table 1, note that a negative number means more diagnoses than expected).

Diagnoses in 2021 among Hispanic/Latino PWH and males (at birth) showed a rebound close to, but still lower levels than, pre-COVID diagnosis levels. This indicates that while the diagnosis deficit from 2020 was not further exacerbated, no significant progress had been made towards closing this deficit in those subpopulations (Table 1).

Diagnosis levels in 2021 among all remaining populations - Black PWH, PWH living in the West and Midwest, females, heterosexual persons, PWIDs, and MSM/PWIDs – rebounded somewhat but remained lower than expected in 2021. As a result, the COVID-related cumulative diagnosis deficit increased among these subpopulations when including 2021. Plots showing percentage of diagnoses missed (or recovered) with respect to expectation in 2020 and 2021, for each subpopulation, are provided in the supplemental figures section at the end of this document.

## Discussion

Our estimates show that more HIV diagnoses occurred in 2021 than expected, among those infected between 2011-2019. While diagnoses appear to have returned to pre-COVID levels in 2021, little progress was made towards eliminating the diagnosis deficit from 2020. A return to the pre-COVID diagnosis level is insufficient as diagnoses must exceed pre-COVID trends to offset the deficits resulting from COVID. Overall, roughly 3,090 HIV diagnoses, from infections prior to 2020, that would have normally been identified remained unidentified by the end of 2021.

In all populations examined, there were fewer missed diagnoses in 2021 than in 2020, suggesting some level of recovery in identifying HIV diagnoses in 2021 after substantial COVID setbacks in 2020. However, the extent of recovery varied by population. Among White PWH, MSM, and PWH in the South and Northeast, diagnosis levels in 2021 were higher-than-expected, where estimated missed diagnoses were negative. This is encouraging and does indicate progress towards reducing the diagnosis deficit among these groups. However, these recoveries were substantially smaller than the corresponding drops in 2020, and significant deficits remain. In the remaining subpopulations considered, the level of recovery was more limited as the estimated number of missed diagnoses in 2021 remained greater than zero. This means that the COVID-related diagnosis deficit in 2020 grew even larger by the end of 2021. More efforts need to be made to expand testing programs to offset these growing

We emphasize that this analysis does not account for infections acquired after 2019, or before 2011. These findings therefore have quantitative significance only when discussing infections acquired over these years. However, they provide qualitative significance in assessing the larger trends in HIV diagnosis data. More concretely, COVID-19 may have changed levels HIV transmission behavior significantly. Determining to what extent an observed change in HIV diagnosis levels may be explained by changes in testing, versus changes in transmission behavior, is not straightforward in general. This cohort examined HIV infections acquired before the onset of the COVID-19 pandemic, so the observed changes in diagnosis levels are independent of any changes in HIV incidence. Our findings are useful in analyzing whether, to what extent, and among which populations, COVID-19 continued to affect HIV diagnoses in 2021, independently of any changes in HIV incidence.

In summary, our findings suggest that insufficient progress was made in 2021 towards reducing the substantial diagnosis deficits incurred in 2020. This is a matter of serious public health concern, as PWH with undiagnosed, and consequently, untreated infection are substantially more likely to transmit HIV [6-8]. A continued, persistent gap in diagnosis levels may serve to further compound this problem, and interventions focused on identifying PWH who have undiagnosed infection are critical. Large deficits persist in all examined subpopulations, as of the end of 2021. Even among the groups that showed higher-than-expected levels of diagnoses in 2021, these increases were substantially smaller than the corresponding decreases observed in 2020. Both general and focused interventions are necessary to close these gaps to avoid future increases in incidence.

## Data Availability

All data used is from the National HIV Surveillance System (NHSS). Inquire with authors regarding data availability.

---

**Supplementary Figures:**
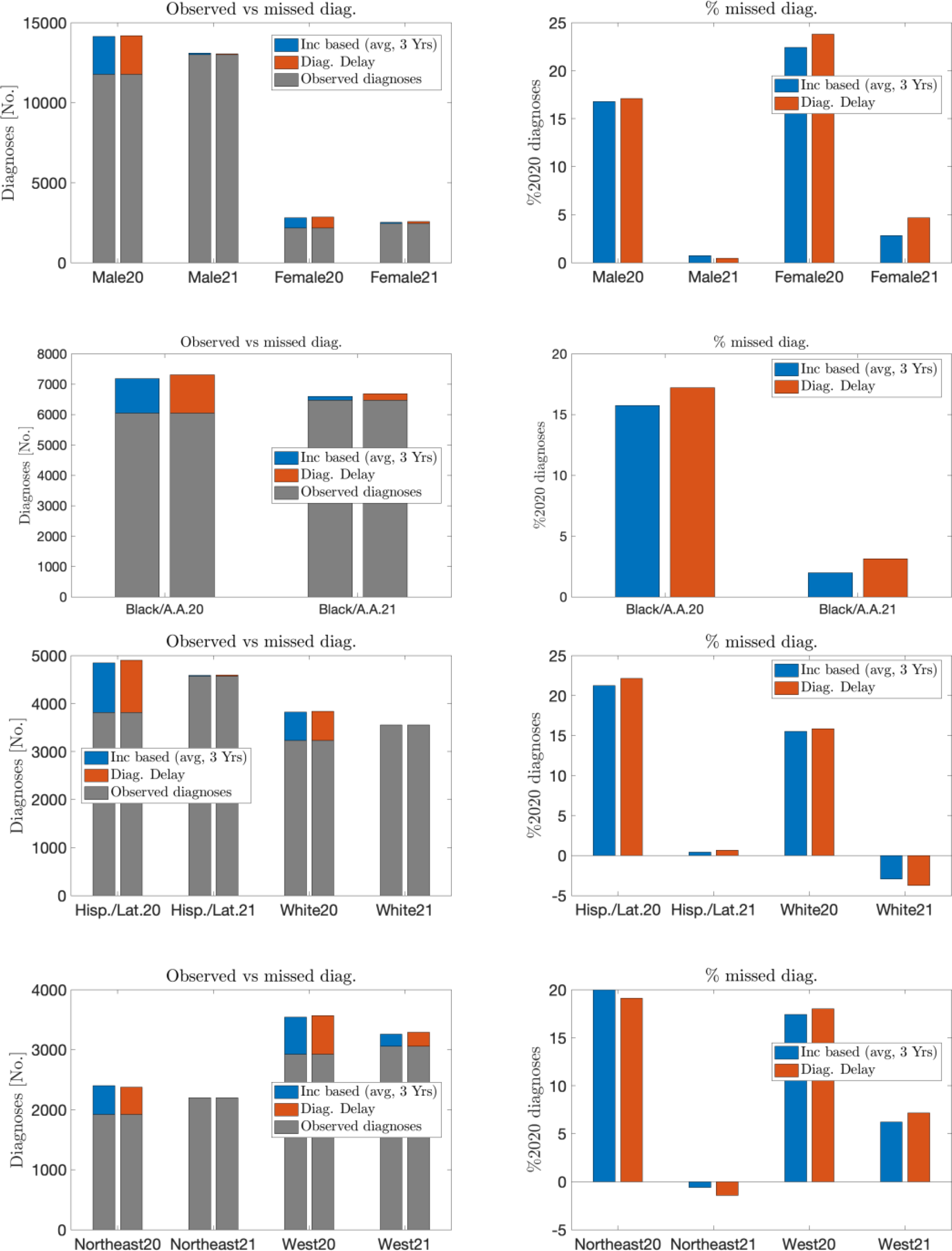
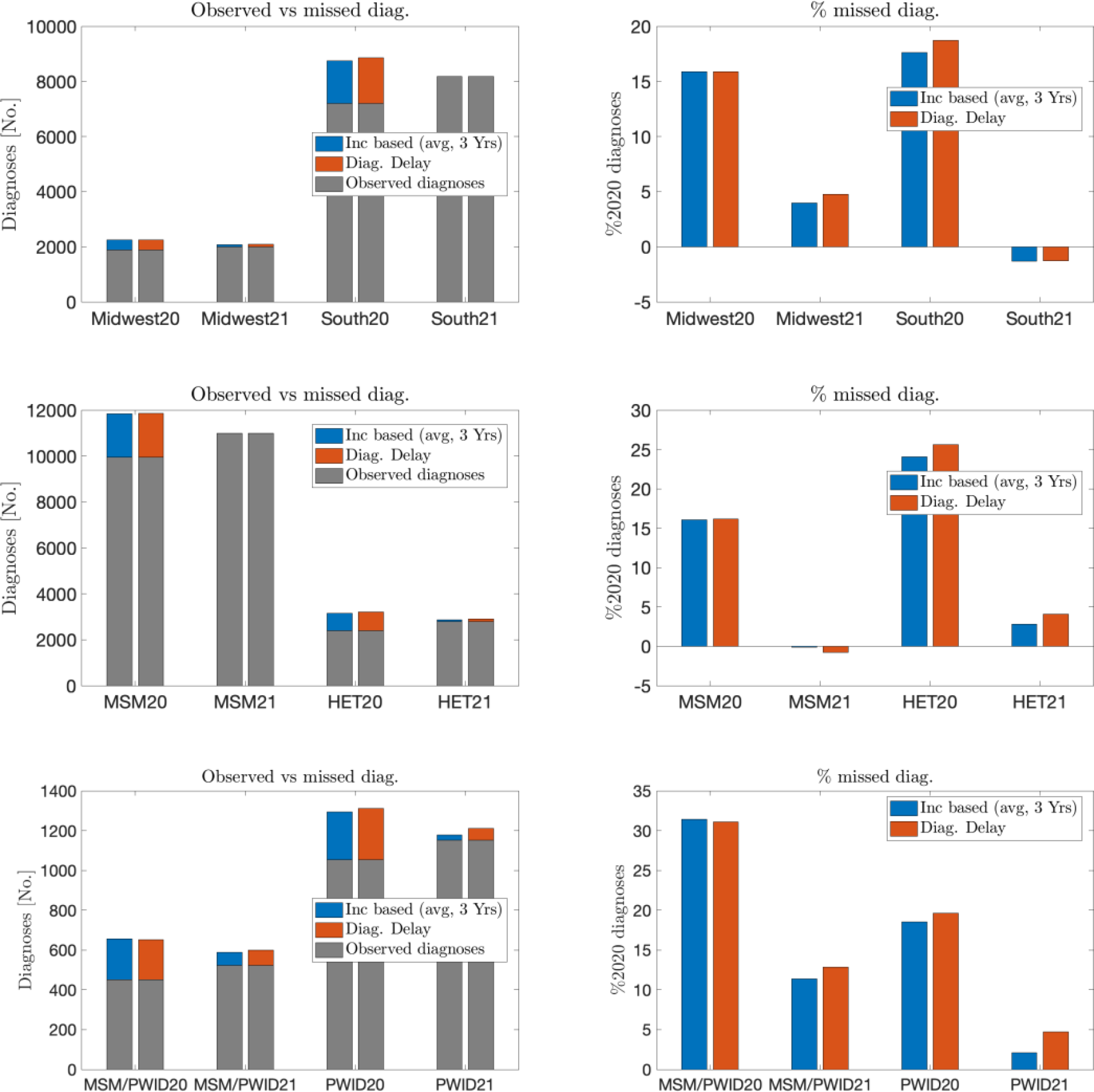
Missed and recovered diagnoses by subpopulation.

## Notes

All support for this project was provided by the Centers for Disease Control and Prevention.

### Competing Interest Statement

The authors have declared no competing interest.

### Funding Statement

Centers for Disease Control and Prevention

## References

[1] E. A. DiNenno et al., “HIV Testing Before and During the COVID-19 Pandemic - United States, 2019-2020,” MMWR Morb Mortal Wkly Rep, vol. 71, no. 25, pp. 820–824, 2022.

[2] “HIV Surveillance Report, 2020,” Centers for Disease Control and Prevention, 2022, vol. 33. Accessed: 6/1/2022. [Online]. Available: https://www.cdc.gov/hiv/library/reports/hiv-surveillance.html

[3] A. Viguerie, R. Song, A. J. Satcher Johnson, C. M. Lyles, A. Hernandez, and P. G. Farnham, “Isolating the Effect of COVID-19-Related Disruptions on HIV Diagnoses in the United States in 2020,” JAIDS, vol. 92, no. 4, pp. 293-299, 2023, doi: 10.1097/QAI.0000000000003140.

[4] R. Song, H. I. Hall, T. A. Green, C. L. Szwarcwald, and N. Pantazis, “Using CD4 Data to Estimate HIV Incidence, Prevalence, and Percent of Undiagnosed Infections in the United States,” JAIDS, vol. 74, no. 1, pp. 3–9, 2017, doi: 10.1097/QAI.0000000000001151.

[5] “HIV Surveillance Report, 2021,” Centers for Disease Control and Prevention, 2023, vol. 34. [Online]. Available: http://www.cdc.gov/hiv/library/reports/hiv-surveillance.html

[6] Q. Xia et al., “Estimating the probability of diagnosis within 1 year of HIV acquisition,” AIDS, vol. 34, no. 7, pp. 1075–1080, 2020, doi: 10.1097/QAD.0000000000002510.

[7] J. Skarbinski et al., “Human Immunodeficiency Virus Transmission at Each Step of the Care Continuum in the United States,” JAMA Intern Med, vol. 175, no. 4, p. 588, 2015, doi: 10.1001/jamainternmed.2014.8180.

[8] Z. Li, D. W. Purcell, S. L. Sansom, D. Hayes, and H. I. Hall, “Vital Signs: HIV Transmission Along the Continuum of Care — United States, 2016,” MMWR Morb Mortal Wkly Rep, vol. 68, no. 11, pp. 267–272, 2019, doi: 10.15585/mmwr.mm6811e1.

